# Covariate-Adjusted Allelic Inference on the Non-Pseudoautosomal X Chromosome with Application to Sex Differences in Allele Frequency

**DOI:** 10.64898/2026.11.25.26368972

**Authors:** Desmond Zeya Chen, Clement Ma

**Affiliations:** Biostatistics Core, Centre for Addiction and Mental Health, Toronto, ON M6J 1H4, Canada; Division of Biostatistics, Dalla Lana School of Public Health, University of Toronto, Toronto, ON M5T 3M7, Canada

**Keywords:** allele frequency, sex differences, covariate adjustment, retrospective regression, X chromosome, heteroskedasticity

## Abstract

Sex differences in minor allele frequency (sdMAF) are widely used as a quality control (QC) metric for variants in the non-pseudoautosomal region (NPR) of the X chromosome. However, conventional sdMAF tests assess marginal allele-frequency differences and generally do not account for imbalances in measured covariates, such as genetic ancestry. We develop a general regression-based framework for covariate-adjusted subgroup allele-frequency inference. Using the Frisch-Waugh-Lovell theorem, we derive adjusted subgroup mean estimators and their joint sampling covariance under a groupwise residual-variance structure, enabling both targeted one-degree-of-freedom tests and joint tests of multiple subgroup contrasts. For X-NPR sdMAF analysis, we consider a parsimonious groupwise variance estimator motivated by the distinct genotype distributions of hemizygous males and diploid females, alongside the conventional homoskedastic and heteroskedasticity-robust HC1 estimators. Simulation studies evaluate Type I error control, robustness to Hardy-Weinberg disequilibrium, and statistical power. The homoskedastic variance estimator exhibited Type I error distortion under imbalanced sex ratios, whereas HC1 and the groupwise estimator demonstrated broadly comparable Type I error control and power across the simulated scenarios. In an application to 1000 Genomes Project data, adjusted and unadjusted sdMAF estimates were highly correlated, although covariate adjustment altered effect estimates and statistical significance for individual variants. The framework also supports a theoretical extension to sex-prevalence-adjusted allelic contrasts, with groupwise variance estimation providing a parsimonious alternative to HC1 for X-NPR sdMAF analysis.

## 1 Introduction

Sex differences in genetic variation have important implications for population genetics, genetic association studies, and genetic data quality control (QC). In particular, sex differences in minor allele frequency (sdMAF) have been used as a QC metric for variants in the non-pseudoautosomal region (NPR) of the X chromosome, where males are hemizygous and females are diploid. Significant sdMAF has been reported in the 1000 Genomes Project, gnomAD, and UK Biobank, motivating closer examination of both technical and biological sources of these differences [1, 2]. The sdMAF test evaluates the null hypothesis of equal allele frequencies between males and females. Significant sdMAF may indicate technical artifacts, including sequencing or genotyping errors, differential missingness, and sex-specific QC procedures. However, observed sdMAF does not necessarily imply poor data quality, as allele-frequency differences may also arise from evolutionary processes or selection associated with disease status and study participation. These considerations highlight the importance of accounting for relevant covariates when interpreting and testing sdMAF.

Conventional sdMAF tests compare allele frequencies between males and females using allele-count-based tests, with variance estimation accounting for their distinct genotype distributions and potential departures from Hardy-Weinberg equilibrium in diploid genotypes [1, 3]. However, these tests assess marginal allele-frequency differences, which may be influenced by imbalances in covariate distributions between sexes. For example, when allele frequencies vary across ancestral populations, differences in ancestry composition between male and female samples can produce marginal sdMAF even when allele frequencies are identical within each ancestral population. This motivates inference on covariate-adjusted allele-frequency differences, a target distinct from the marginal contrast. Regression provides a natural approach to such adjustment. Adjusted subgroup means share nuisance coefficients estimated from the full sample, introducing uncertainty and potentially nonzero covariance among the adjusted estimates. Inference on their contrasts must therefore account for this joint covariance. Moreover, hemizygous male and diploid female genotypes in the X-NPR exhibit different variance-mean relationships even when allele counts are placed on a common scale, so a common residual-variance assumption need not hold.

Regression-based allelic methods provide a foundation for covariate adjustment. Zhang and Sun developed a robust allele-based score test for autosomal association analysis that accommodates covariates and departures from HWE [4]. For sdMAF, Wang et al. proposed a population-aware retrospective regression with sex-by-population interactions and group-specific residual variances. Although their model permits additional covariates, these were omitted from the explicit test derivations [5]. More recently, Zhang et al. developed an X-chromosomal HWE framework that distinguishes female HWD from sdMAF, with the derived X-NPR tests formulated without additional nuisance covariates [6]. These developments motivate an explicit treatment of covariate-adjusted X-NPR allele-frequency contrasts, their joint sampling covariance, and the residual-variance assumptions required for inference.

We develop a covariate-adjusted inferential framework for assessing subgroup differences in allele frequency, with a particular focus on sdMAF in the X-NPR. We define adjusted subgroup mean estimators, express them using the Frisch-Waugh-Lovell theorem, and derive their joint sampling covariance under a groupwise residual-variance structure. These results provide estimates, standard errors, and tests for prespecified subgroup allele-frequency contrasts, with covariate-adjusted sdMAF as a two-group application. Through simulation studies, we evaluate the groupwise variance estimator against the conventional homoskedastic estimator, the heteroskedasticity-robust Huber-White HC1 estimator, and the unadjusted one-degree-of-freedom sdMAF test in terms of Type I error control and statistical power. Finally, we apply the proposed framework and comparator methods to data from the 1000 Genomes Project to investigate the impact of covariate adjustment on sdMAF inference in the X-NPR. Although the framework accommodates more general subgroup allele-frequency contrasts, our empirical investigations focus on sdMAF.

## 2 Methods

### 2.1 General regression formulation

Let *G*_*i*_ denote the genotype at a bi-allelic SNP, typically coded additively as the number of minor alleles, *G*_*i*_ ∈ {0, 1, 2}, which applies to all females genome-wide and to males for autosomal and pseudoautosomal (PAR) variants. For variants in the X chromosome non-pseudoautosomal region (NPR), hemizygous males are coded as *G*_*i*_ ∈ {0, 2} to place genotypes on a common allele-frequency scale across sexes.

Let *S*_*i*_ ∈ {*f, m*} denote biological sex, with *f* = female *≡* 0 and *m* = male *≡* 1. Let **Z**_*i*_ = (*Z*_*i*1_, …, *Z*_*iq*_)^T^ denote a set of additional baseline covariates, such as genetic principal components or age, with corresponding coefficient vector ***γ*** = (*γ*_1_, …, *γ*_*q*_)^T^. Write **Z** = (**Z**_1_, …, **Z**_*n*_)^T^ and **G** = (*G*_1_, …, *G*_*n*_)^T^.

Consider the linear model, **G** = **X*β*** + **Z*γ*** + ***ε***, where **G** ∈ R^*n×*1^, **X** ∈ R^*n×r*^, **Z** ∈ R^*n×q*^ and ***ε*** = (*ε*_1_, …, *ε*_*n*_)^T^ has independent, conditionally mean-zero components with heteroskedasticity. The primary covariates **X**_*i*_ may include categorical variables, continuous variables, their transformations, interaction terms, or any combination of these, with corresponding coefficient vector ***β*** ∈ R^*r×*1^. Assume **H** = [**X, Z**] has full column rank, with *d* = rank(**H**) = *r* + *q < n*. Assume the data has been grouped and ordered for notation simplicity with *f* (**X**_*i*_) = *k*, a functional that groups observation *i* to group *k* ∈ {1, …, *K*} based on its primary covariates **X**_*i*_. We additionally assume the nuisance covariates **Z** are centered. Then the conditional variance of **G** consists of blocks of group specific variance-covariance matrix 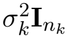.

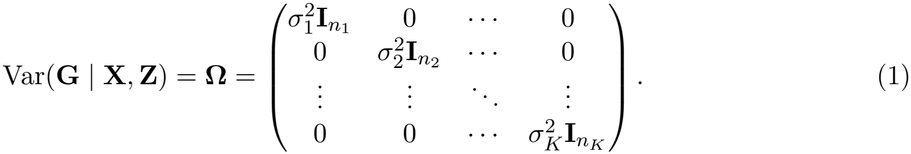

Let *I*_*k*_ := {*i* ∈ {1, …, *n*} : *f* (**X**_*i*_) = *k}* and *n*_*k*_ = |*I*_*k*_|, where *I*_*k*_ denotes the indices of observations belonging to group *k*. Define the selection matrix 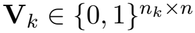 by

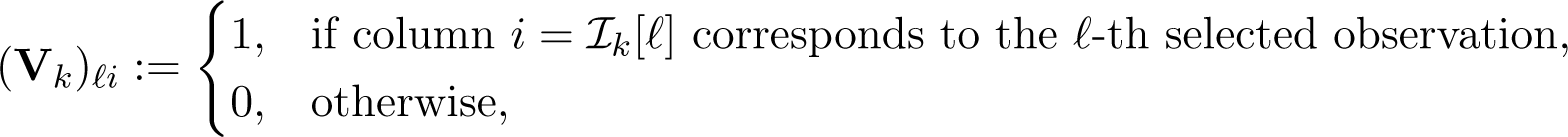

so that the subvector of selected observations can be written as

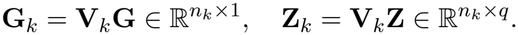

Here, 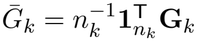 and 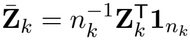, where 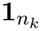 is an *n*_*k*_ *×* 1 vector of ones.

We additionally require each group-indicator vector 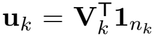 to belong to col(**X**). Thus, **M**_*X*_**u**_*k*_ = **0**. This condition permits additional continuous primary covariates but requires the design to represent each group’s indicator. The intercept is included in **X** and is not duplicated in **Z**.

### Adjusted group means

By the Frisch-Waugh-Lovell theorem [7, 8],

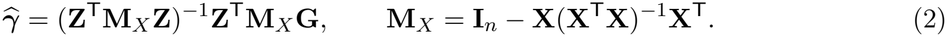

Define

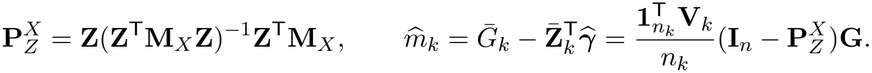

Here 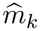 estimates 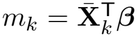, where 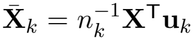. Under the group-indicator condition, the OLS normal equations give 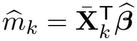.

### Joint covariance of adjusted group means

Under the groupwise variance structure, 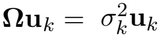. The group-indicator condition gives **M**_*X*_**u**_*k*_ = **0**, and hence 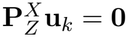.

For a single group, define 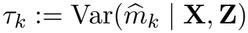. Using the linear representation of 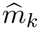, we obtain

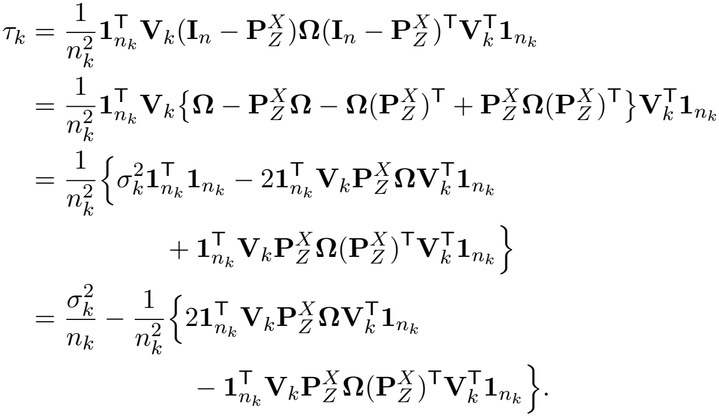

The two cross terms in the expansion are transposes of the same scalar and therefore equal. Moreover,

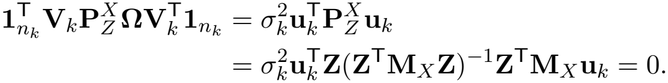

Consequently, the closed-form variance is

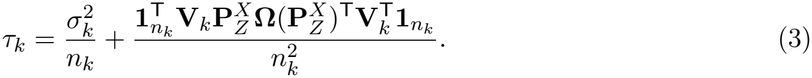

For two distinct groups *k ≠ l*, define 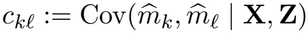. Similarly,

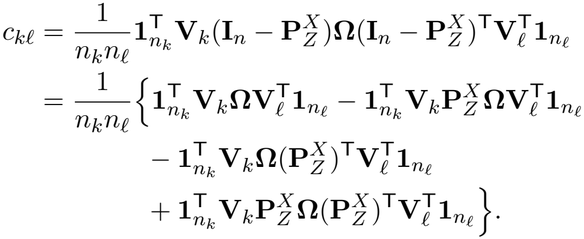

The first term is zero because the groups are disjoint and observations are conditionally independent:

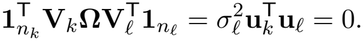

The second and third terms also vanish, since

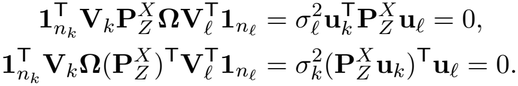

Here the second term uses 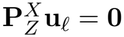, whereas the third uses 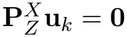. Thus, only the covariance between the two estimated nuisance contributions remains:

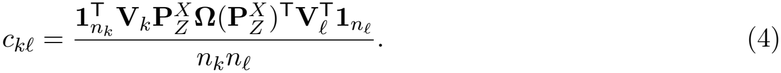

Equivalently, the additional term in *τ*_*k*_ is 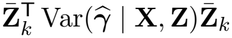, and 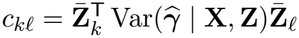, where

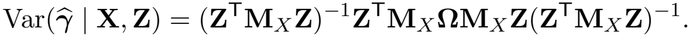

Let 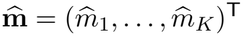. Its conditional covariance matrix is

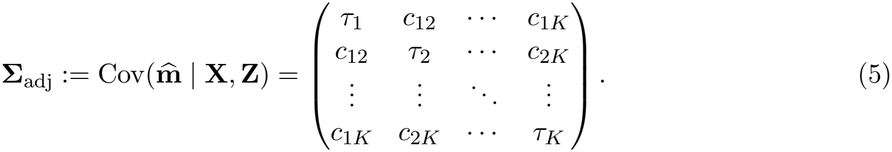

For direct computation, define **L** with row *k* equal to 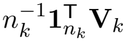. Then

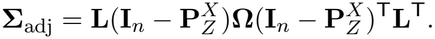

This matrix identity also holds without the group-indicator condition or blockwise-constant residual variances; the simplified elements in equations (3)-(4) use both conditions.

For estimation, let 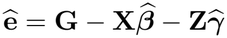 and 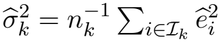. We use

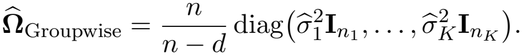

The plug-in covariance 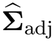 is obtained from the full matrix identity above. If the simplified elements are used, the same correction *n/*(*n −d*) is applied to all occurrences of the estimated residual variances. Alternative residual covariance estimators, including HC1, can also be substituted into the full matrix identity.

#### Linear contrasts and joint Wald tests

Let **m** = (*m*_1_, …, *m*_*K*_)^T^ and let **C** *∈*ℝ^*h×K*^ be a prespecified contrast matrix with full row rank *h*. For testing *H*_0_ : **Cm** = **0**, the Wald statistic is

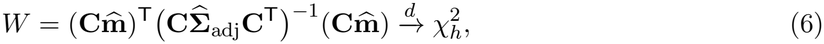

under the null and the usual consistency and asymptotic-normality conditions, with nonsingular contrast covariance. For contrasts of group differences, **C1**_*K*_ = **0**. A pairwise difference uses a single row with entries 1 and *−*1 at the two selected groups, giving estimated variance 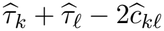.

### 2.2 Application to X chromosome sdMAF

For a common effect allele at a biallelic X-NPR variant, code female genotypes as *G*_*i*_ ∈ {0, 1, 2} and male genotypes as *G*_*i*_ ∈ {0, 2}. The marginal effect-allele frequencies are

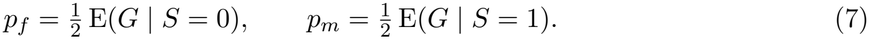

This coding places both sexes on a common allele-frequency scale; it does not impose a biological assumption about X-chromosome inactivation or phenotypic dosage compensation.

Under the common-slope model

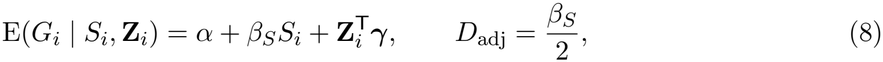

*D*_adj_ is the male-female allele-frequency contrast at a common covariate value. It differs from the marginal contrast *p*_*m*_ *− p*_*f*_ when covariate mean contributions differ between sexes, since *p*_*m*_*− p*_*f*_ = *β*_*S*_*/*2 + (E(**Z** |*S* = 1) *−* E(**Z**| *S* = 0))^T^***γ****/*2. Here **X** = [**1**_*n*_, **S**], ***β*** = (*α, β*_*S*_)^T^ and *d* = 2 + *q*. We use one residual variance group per sex. The equality *K* = *r* = 2 is specific to this application, not a requirement of the general framework.

The adjusted estimator is

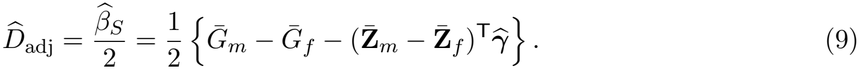

Thus the adjusted two-group comparison and the sex coefficient yield the same estimator and, using the same residual covariance specification, the same estimated variance. With 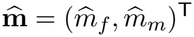, take 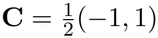 in equation (6). Then

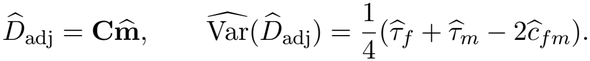

The null *H*_0_ : **Cm** = 0 is equivalent to *H*_0_ : *m*_*m*_ = *m*_*f*_, or *H*_0_ : *β*_*S*_ = 0, and the resulting Wald statistic has an asymptotic 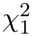 null distribution.

The distinct genotype distributions motivate sex-specific variance estimation. Defining 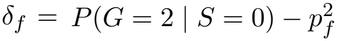 to describe female Hardy-Weinberg disequilibrium gives

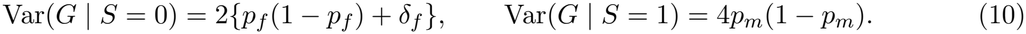

These are marginal genotype variances, not conditional residual variances after adjustment. Groupwise further assumes constant conditional residual variance within each sex; HC1 relaxes this restriction.

#### Variance estimation and testing

Let **e**_*S*_ *∈* ℝ^*d*^ select the sex coefficient in the full model, with row *i* of **H** denoted by 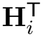. Define

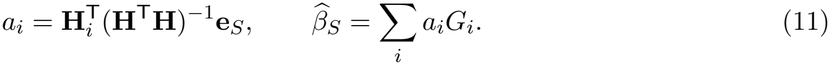

The general covariance estimators reduce to

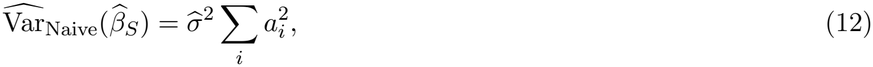

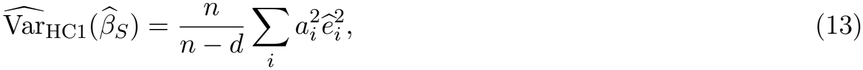

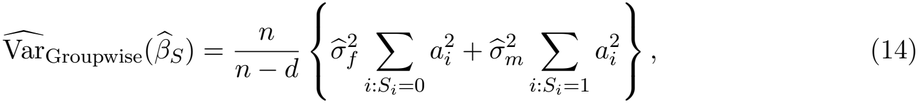

where 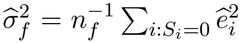 and 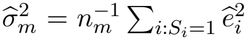. On the allele-frequency scale, 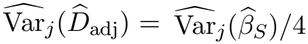. Testing *H*_0_ : *D*_adj_ = 0 is therefore equivalent to testing *H*_0_ : *β*_*S*_ = 0, using

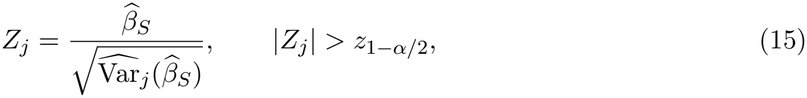

where *z*_1*−α/*2_ is the standard normal quantile. Equivalently, 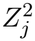 is compared with 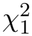. We use Groupwise as the working variance specification and compare it with Naive and HC1 in the sdMAF evaluation. These comparisons concern variance estimation, not different adjusted point estimators.

### 2.3 Extension to SPADE

For a case-control application, let *Y*_*i*_ ∈ {*a, u*} denote case or control status, respectively, and let *π*_*s*_ = *P* (*Y* = *a*| *S* = *s*) denote known sex-specific disease prevalence in the target population. Use the four sex-by-disease groups, ordered as (*f, a*), (*f, u*), (*m, a*), (*m, u*), and a primary design that contains their indicators in its column space, such as an intercept, sex, disease status, and their interaction. Define

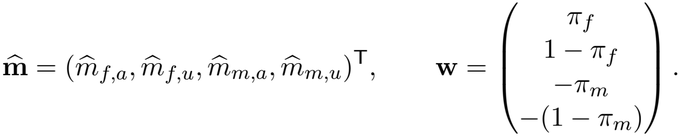

The corresponding covariance matrix is

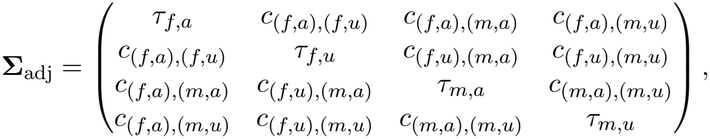

whose elements are given by equations (3)-(4). On the common genotype scale used here, the prevalence-weighted adjusted allele-frequency contrast and its estimated variance are

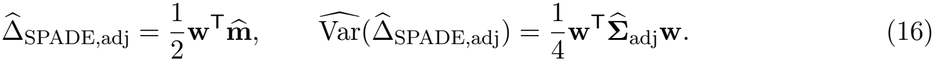

Thus, testing the weighted adjusted contrast against zero is a one-degree-of-freedom instance of equation (6), with **C** = **w**^T^*/*2. The full covariance is retained because the four adjusted means share the estimated nuisance coefficients.

This contrast compares prevalence-weighted group means at the common nuisance-covariate reference **Z** = **0**. It need not equal the unadjusted population allele-frequency difference, which additionally averages over the target population’s covariate distributions. The prevalence parameters are treated as fixed; their estimation uncertainty is not included. This extension is theoretical and is not evaluated in the simulations or real-data analysis below.

### 2.4 Simulation studies and real data application

#### 2.4.1 Simulation setup

We conducted simulation studies to evaluate the finite-sample performance of the proposed covariate-adjusted allelic test for sex differences in allele frequency on the non-pseudoautosomal region (NPR) of the X chromosome. Each simulated dataset consisted of *n* = 5,000 individuals. Five covariates were generated independently as *Z*_1_ ~ Bernoulli(0.4), *Z*_2_ ~ Categorical(0.5, 0.3, 0.2), *Z*_3_ ~ Uniform(0, 1), *Z*_4_, *Z*_5_ *~ N* (0, 0.01^2^). The three levels of *Z*_2_, coded 0, 1, and 2, were represented by two dummy variables, *Z*_2*B*_ = *I*(*Z*_2_ = 1) and *Z*_2*C*_ = *I*(*Z*_2_ = 2).

Sex was generated conditional on the covariates. Let *S*_*i*_ = 1 denote a male individual and *S*_*i*_ = 0 denote a female individual. We first defined

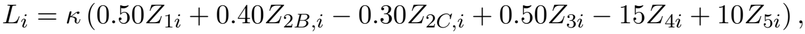

where *k* controls the overall strength of the covariate effects. The covariate score was centered within each simulated dataset as

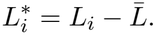

Sex was then generated according to

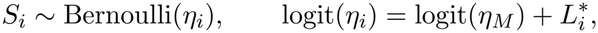

where *η*_*M*_ is the male-proportion parameter used in the logistic intercept. Centering the score on the logit scale does not force the marginal male probability to equal *η*_*M*_; the observed male proportion was recorded across replicates.

Conditional on sex and the covariates, the individual-specific allele frequency was defined as

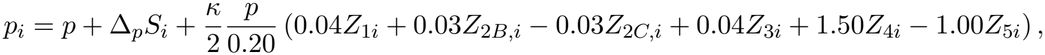

where *p* is the baseline MAF and Δ_*p*_ is the conditional male-female difference in allele frequency. Thus, the same covariates affect both sex and allele frequency when *k >* 0, creating covariate-induced differences in the marginal allele-frequency distributions between males and females even when Δ_*p*_ = 0. The factor *p/*0.20 scales the magnitude of the covariate effects according to the baseline MAF.

Female genotypes were generated allowing for Hardy-Weinberg disequilibrium (HWD). Let *A*_*i*_ denote an indicator of whether female individual *i* was subject to HWD. We defined

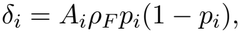

where *ρ*_*F*_ controls the magnitude and direction of departure from Hardy-Weinberg equilibrium. Conditional on the individual-specific allele frequency *p*_*i*_, the female genotype probabilities were

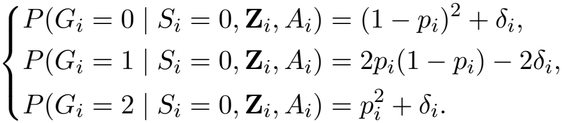

This parameterization preserves the conditional mean,

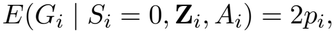

while allowing the conditional variance to depart from that under Hardy-Weinberg equilibrium:

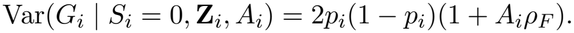

When *A*_*i*_ = 0 or *ρ*_*F*_ = 0, genotypes are generated under Hardy-Weinberg equilibrium. When *A*_*i*_ = 1, *ρ*_*F*_ *<* 0 corresponds to heterozygote excess and *ρ*_*F*_ *>* 0 to heterozygote deficiency.

Male hemizygous genotypes on the X-chromosome NPR were generated as

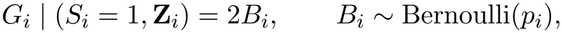

such that

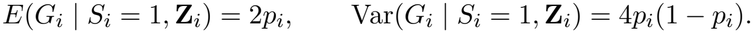

Thus, the genotype-generating mechanism preserves the common conditional mean

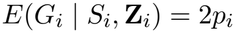

across sexes while allowing sex-specific conditional genotype variances. When *ρ*_*F*_ = 0, female genotypes satisfy Hardy-Weinberg equilibrium conditional on their covariates, whereas *ρ*_*F*_ *>* 0 induces excess homozygosity and a corresponding reduction in heterozygosity.

For covariate-adjusted analyses, we fitted the linear model

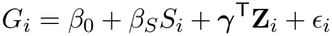

and tested

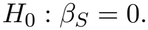

Under the specified data-generating model,

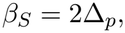

such that 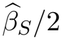 estimates the covariate-adjusted difference in allele frequency between males and females. We considered three variance estimators for 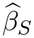 the conventional homoskedastic variance estimator (Naive), the HC1 heteroskedasticity-consistent variance estimator (HC1) [9, 10], and a groupwise estimator allowing separate residual variances for males and females (Groupwise). These approaches were additionally compared with the conventional unadjusted sdMAF test based directly on the observed male and female allele frequencies.

The genotype-generating covariates were used uncentered in the simulation code; centering them in the fitted model only changes the intercept, not the estimated sex coefficient or its variance. The adjusted design contained an intercept, sex, and six nuisance columns, giving *d* = 8. The baseline parameter *p* is the allele frequency at *S* = 0 and zero covariates, not necessarily the sample-wide MAF. Individual probabilities were checked to satisfy 0 *< p*_*i*_ *<* 1; the code stopped on an invalid value rather than truncating or resampling it.

Although the mean model is correctly specified, conditional residual variances depend on *p*_*i*_ and therefore generally vary with covariates within each sex. These simulations consequently assess the adequacy of the working sex-groupwise variance estimator beyond an exactly constant within-sex variance structure. Conditional HWE also does not imply HWE after pooling individuals with different *p*_*i*_.

#### 2.4.2 Type I error and HWD sensitivity

Type I error was first evaluated under the null hypothesis of no conditional sex difference in allele frequency by setting

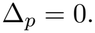

For the primary simulations, female genotypes were generated under Hardy-Weinberg equilibrium by setting *ρ*_*F*_ = 0. We considered baseline MAFs

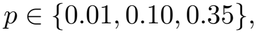

covariate-effect multipliers

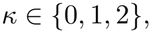

representing no, moderate, and strong covariate effects, respectively, and target male proportions

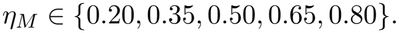

The full factorial design therefore consisted of 3*×* 3*×* 5 = 45 primary simulation scenarios.

We additionally conducted a sensitivity analysis to evaluate the impact of female HWD on test calibration. For these simulations, the baseline MAF and covariate-effect multiplier were fixed at *p* = 0.30 and *κ* = 1, respectively. The magnitude of female HWD was varied according to

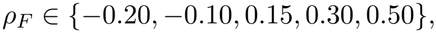

thereby including both heterozygote excess (*ρ*_*F*_ *<* 0) and heterozygote deficiency (*ρ*_*F*_ *>* 0). To allow HWD to affect only a subset of females, an HWD indicator was randomly assigned among female individuals, with the proportion of females subject to HWD set to

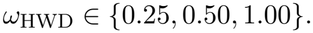

For females not assigned to the HWD subgroup, *ρ*_*F*_ was set to zero. Target male proportions of

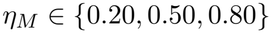

were considered. This resulted in an additional 5 *×*3 *×*3 = 45 HWD sensitivity scenarios. Genotype probabilities were required to remain within the valid probability space for all simulated individuals.

For each scenario, 10,000 independent Monte Carlo replicates of *n* = 5,000 individuals were generated. Four methods were evaluated: the conventional unadjusted sdMAF test, covariate-adjusted regression using the conventional homoskedastic variance estimator (Naive), covariate-adjusted regression using the HC1 heteroskedasticity-consistent variance estimator (HC1), and covariate-adjusted regression using the sex-groupwise residual variance estimator (Groupwise). Each method was evaluated using a two-sided test at the nominal significance level *α* = 0.05. For the adjusted methods, empirical type I error was calculated as the proportion of replicates rejecting the adjusted null. The unadjusted procedure tests a marginal null, which need not hold when *κ >* 0 even if Δ_*p*_ = 0; its rejection rate in those settings was not interpreted as its own Type I error. With 10,000 replicates, the approximate 95% Monte Carlo interval around a true type I error rate of 0.05 was 0.0457-0.0543. We additionally assessed variance estimation by comparing the mean estimated variance with the empirical variance of the corresponding effect estimator across Monte Carlo replicates.

#### 2.4.3 Power

Power was evaluated under alternatives defined by Δ_*p*_ = *λp*, where *λ* denotes the effect multiplier. Thus, the true adjusted sex coefficient was *β*_*S*_ = 2*λp*, rather than 2*λ*. We considered

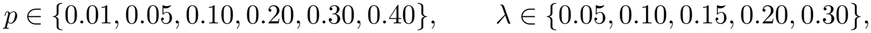

and target male-proportion parameters *η*_*M*_ ∈ {0.20, 0.50, 0.80}, giving 6 *×*5 *×*3 = 90 scenarios. The covariate-effect multiplier was fixed at *κ* = 1. Female genotypes were generated under conditional Hardy-Weinberg equilibrium by setting *ρ*_*F*_ = 0; male genotypes retained the 0/2 coding. The remaining data-generating mechanisms were unchanged from the simulation setup above.

For each scenario, we generated 10,000 Monte Carlo replicates of *n* = 5,000 individuals. Naive, HC1, and Groupwise used the same adjusted OLS sex-coefficient estimate and differed only in variance estimation. Adjusted tests rejected when 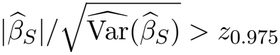. The unadjusted comparator used its one-degree-of-freedom Wald test of the marginal male-female allele-frequency difference. Empirical rejection rates were calculated at *α* = 0.05. Because the unadjusted and adjusted procedures target marginal and conditional contrasts, respectively, their rejection rates need not represent power against the same effect parameter. Power comparisons among adjusted methods were interpreted together with their null calibration.

We also recorded the mean estimated sex coefficient, the empirical variance across replicates, the mean estimated variance, and the ratio of mean estimated to empirical variance. For the unadjusted method, the allele-frequency difference was multiplied by two and its variance by four to express these summaries on the genotype-coefficient scale. The Monte Carlo standard error of a rejection-rate estimate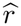 was 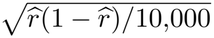 at most 0.005. Random-number seeds were set to 123 + *j* for scenario *j* = 1, …, 90 in the factorial grid.

#### 2.4.4 Application to the 1000 Genomes Project

We applied the proposed covariate-adjusted allelic testing framework to chromosome X data from the high-coverage 1000 Genomes Project. Phased genotype data were obtained from the 20220422 release of the 1000G high-coverage SNV/INDEL/SV dataset, aligned to the GRCh38 reference genome (see Data availability). Variants were left-aligned and normalized against the GRCh38 reference genome, atomized, and split at multiallelic sites using bcftools. The dataset was subsequently restricted to 2,504 individuals from the 1000 Genomes reference panel [11].

To account for population structure, the top 20 genetic ancestry principal components (PCs) for these 2,504 individuals were obtained from the PCs_1000G dataset in the PCAmatchR R package and included as covariates [12]. For comparisons between adjusted and unadjusted analyses, we restricted the data to biallelic single-nucleotide variants (SNVs). Variants were matched by chromosome, genomic position, and allele pair, with ambiguous duplicate matches excluded. Allele orientations were harmonized to ensure that adjusted and unadjusted allelic differences were expressed relative to the same allele. Only variants successfully matched between the two analyses were retained for comparison.

## 3 Results

### 3.1 Type I error under varying sample composition

Across the simulation scenarios, we observed several distinct patterns in empirical Type I error rates (Figure 1). First, the unadjusted test exhibited increasingly inflated Type I error rates with increasing MAF under moderate and strong covariate effects, highlighting the consequences of failing to account for covariate effects. Second, the naive variance estimator showed a systematic dependence on the sex composition of the sample, with inflated rejection rates when males were underrepresented (20% or 35%) and conservative rejection rates when males were overrepresented (65% or 80%). This suggests that the direction of Type I error distortion under the naive variance estimator is sensitive to sex imbalance. In contrast, both the HC1 and groupwise variance estimators maintained empirical Type I error rates within the 95% Monte Carlo reference bounds across nearly all scenarios. The only exceptions occurred at MAF = 0.01 with 20% male participants under no or moderate covariate effects. These isolated departures may reflect finite-sample limitations of the asymptotic Wald approximation when sex-specific minor allele counts are particularly small.

**Figure 1:**
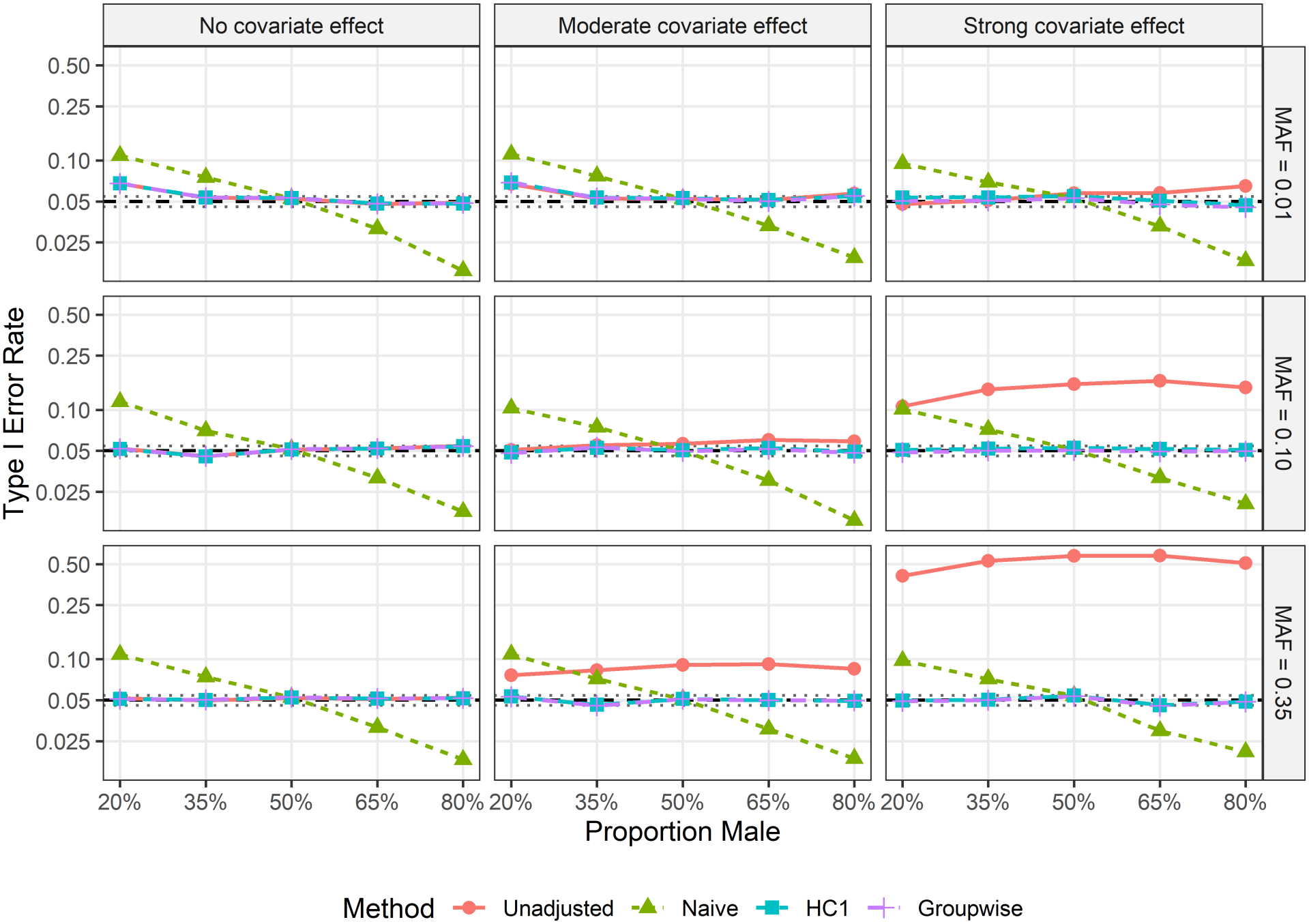
Empirical Type I error rates across simulation scenarios. Each scenario consisted of 10,000 Monte Carlo replicates with a sample size of 5,000 and a nominal significance level of 0.05. Four methods were evaluated: Unadjusted (without covariate adjustment), Naive (covariate-adjusted with the conventional homoskedastic variance estimator), HC1 (covariate-adjusted with the HC1 heteroskedasticity-consistent variance estimator), and Groupwise (covariate-adjusted with the proposed groupwise variance estimator). The dashed horizontal line indicates the nominal Type I error rate, while the 95% Monte Carlo reference bounds (0.0457-0.0543) indicate the expected range of empirical rejection rates under the null hypothesis due to simulation variability. Estimates above the upper bound suggest potential Type I error inflation, whereas estimates below the lower bound suggest conservativeness.

We further evaluated the sensitivity of empirical Type I error rates to HWD among females (Figure 2). The unadjusted test consistently exhibited inflated Type I error rates across all scenarios, although the magnitude of inflation varied with the simulation settings. Similar to the findings under Hardy-Weinberg equilibrium, the naive variance estimator exhibited a systematic dependence on sex composition, with inflation when males were underrepresented and conservativeness when males were overrepresented. Interestingly, as the magnitude of female HWD increased, the rejection rates of the naive estimator tended to approach the nominal significance level from either direction. The rate of this convergence depended on the proportion of females exhibiting HWD, with a greater proportion associated with a more pronounced attenuation of Type I error distortion. In contrast, HC1 and the groupwise variance estimator demonstrated similar operating characteristics across the HWD scenarios, with no clear systematic divergence between the two methods.

**Figure 2:**
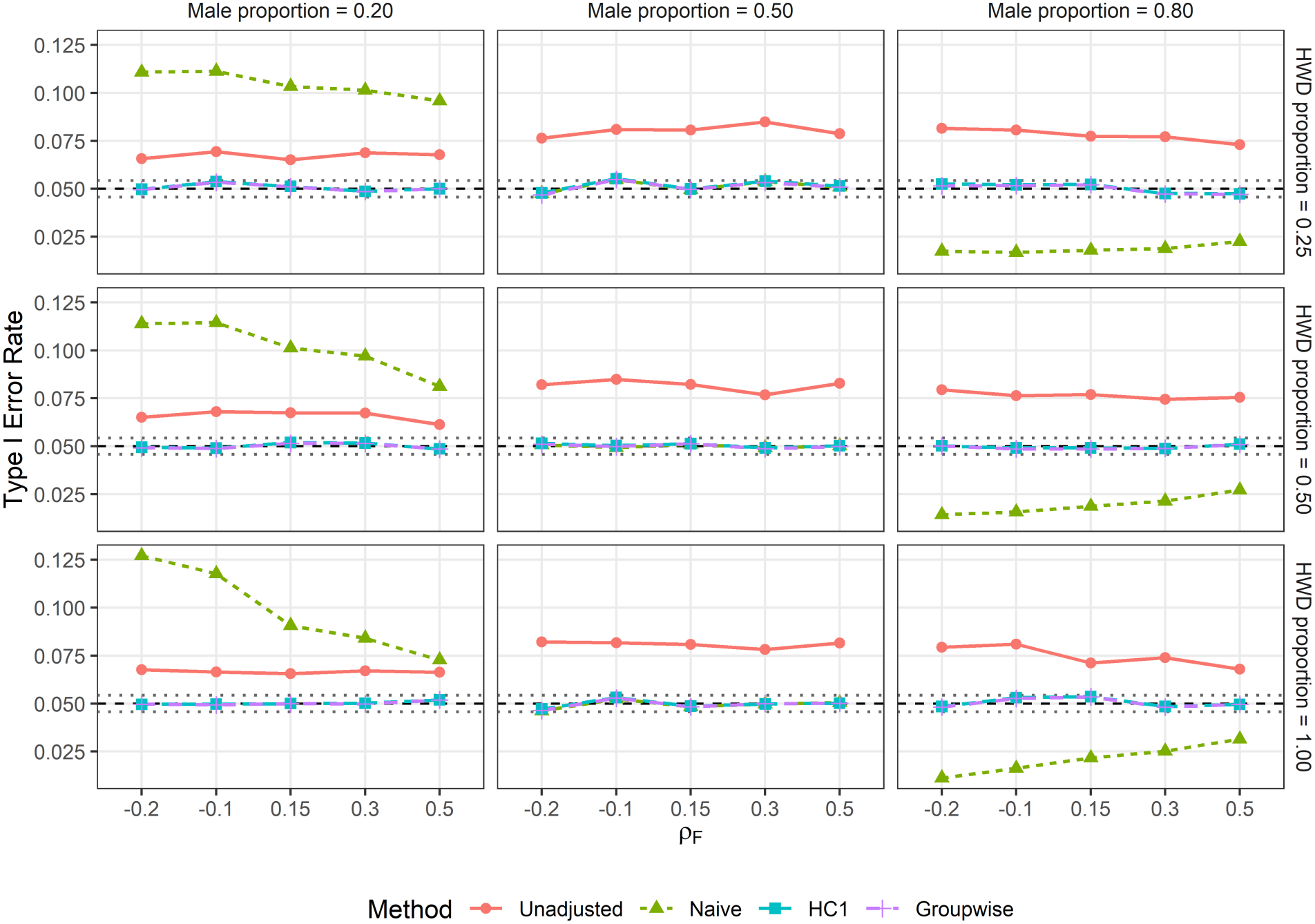
Empirical Type I error rates under varying degrees of female HWD. Each scenario consisted of 10,000 Monte Carlo replicates with a sample size of 5,000 and a nominal significance level of 0.05. The parameter *ρ*_*F*_ represents the degree of HWD among females. Method labels are defined as in Figure 1. The dashed horizontal line indicates the nominal Type I error rate, while the pointwise 95% Monte Carlo reference bounds (0.0457-0.0543) represent the expected range of empirical rejection rates under the null hypothesis due to simulation variability. Values above the upper bound suggest Type I error inflation, whereas values below the lower bound suggest conservativeness.

### 3.2 Statistical power

Empirical statistical power generally increased with increasing minor allele frequency (MAF) and effect size (Figure 3). Consistent with the Type I error simulation results, the unadjusted test exhibited higher empirical rejection rates than the covariate-adjusted methods across most scenarios. However, this apparent power advantage should be interpreted cautiously, as the unadjusted test also exhibited inflated Type I error rates, suggesting that its higher rejection rates may partly reflect inadequate calibration rather than greater statistical efficiency.

**Figure 3:**
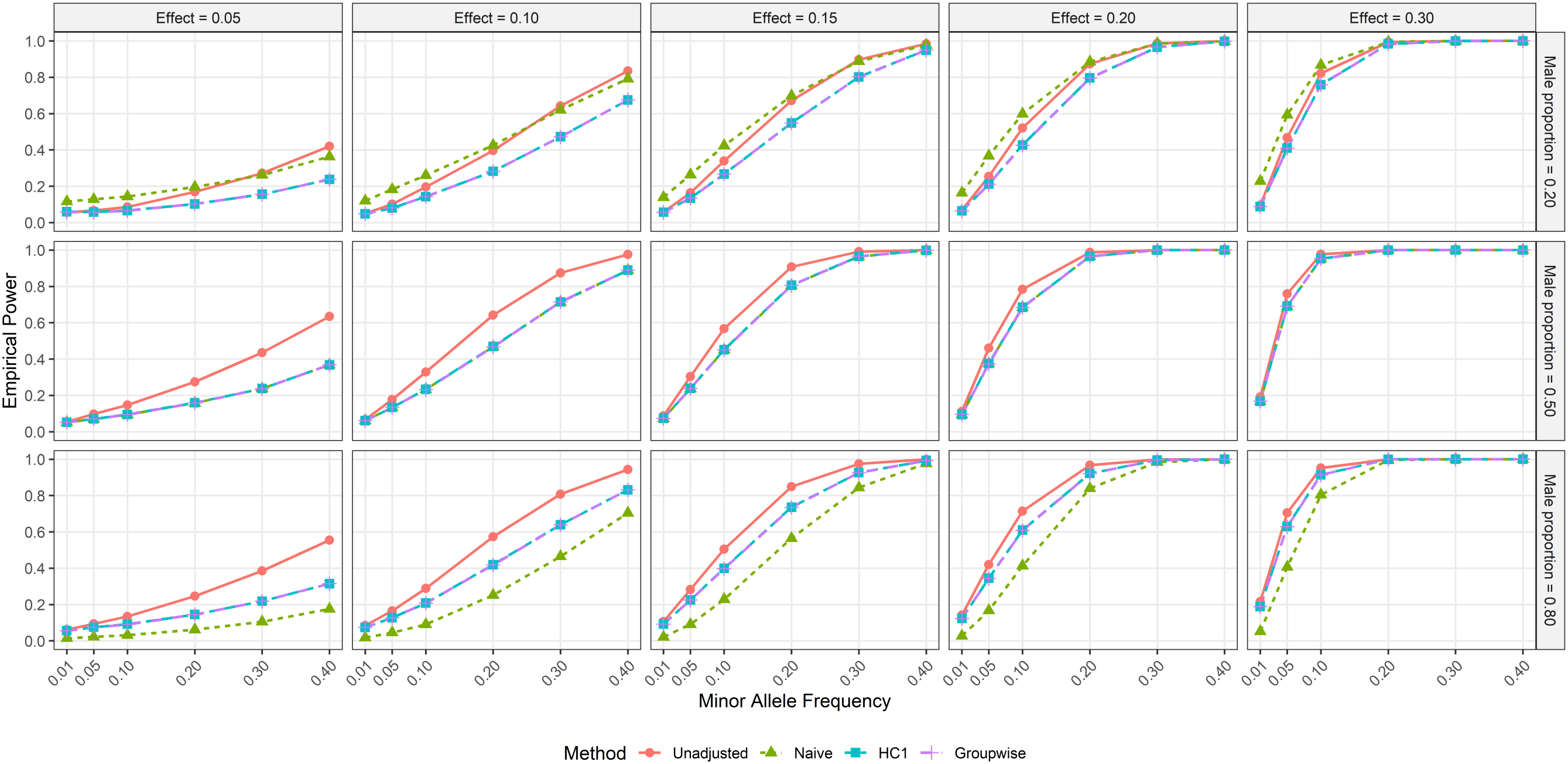
Empirical statistical power across minor allele frequencies, effect sizes, and sex compositions. Each simulation scenario included 5,000 individuals and 10,000 Monte Carlo replicates. Columns represent effect sizes ranging from 0.05 to 0.30, while rows represent male sample proportions of 20%, 50%, and 80%. Minor allele frequency is displayed on the horizontal axis. The four methods are defined as in Figure 1. Empirical power was calculated as the proportion of simulations rejecting the null hypothesis at the nominal significance level of 0.05.

The naive variance estimator showed a similar dependence on sex composition to that observed in the Type I error simulations. When males accounted for 20% of the sample, the naive estimator tended to be anti-conservative, whereas it became conservative when the male proportion increased to 80%. These differences in calibration were reflected in the corresponding empirical power patterns. In contrast, the HC1 and groupwise variance estimators exhibited nearly identical empirical power across all simulation scenarios, consistent with their comparable Type I error control.

### 3.3 Effects of covariate adjustment on sdMAF inference

Covariate adjustment affected sdMAF estimates differently across MAF strata (Figure 4). For variants with MAF below 0.01 and between 0.01 and 0.05, the density plots showed a predominant upward shift relative to the identity line. This corresponds to larger positive sdMAF estimates and negative estimates closer to zero after adjustment. In contrast, variants with MAF between 0.05 and 0.20 and those with MAF at least 0.20 showed a more balanced distribution above and below the identity line, with no similarly pronounced directional shift. These patterns suggest that the effects of genetic principal component adjustment on sdMAF estimates varied across MAF strata in this dataset.

**Figure 4:**
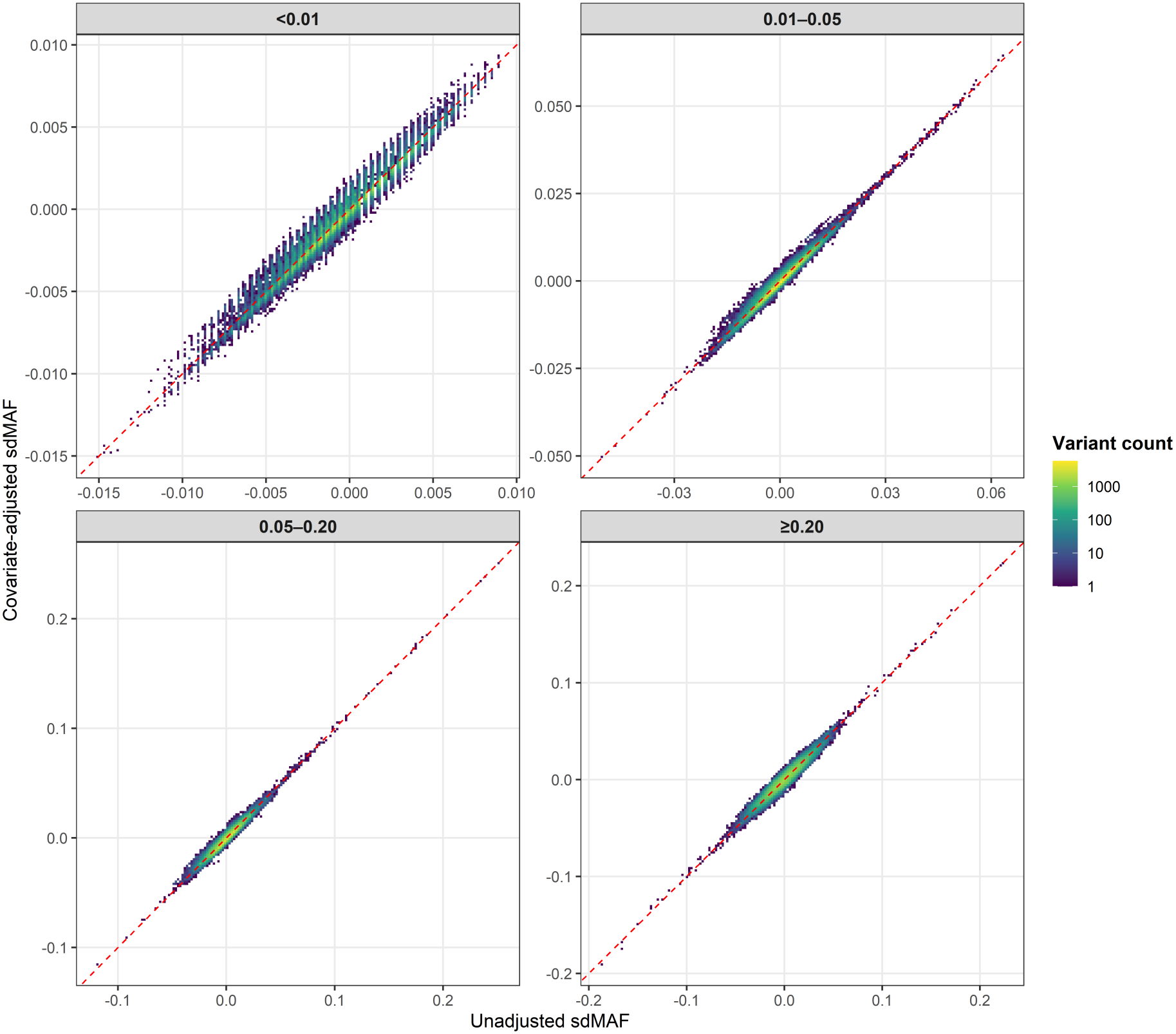
Comparison of unadjusted and covariate-adjusted sdMAF estimates. Two-dimensional binned density plots compare unadjusted estimates (horizontal axis) with covariate-adjusted estimates (vertical axis), stratified into nine MAF categories. Adjusted estimates were obtained from retrospective linear regression and expressed on the allele-frequency scale. Variants were harmonized by genomic position and allele identity. Color indicates the number of variants per bin. The dashed red line denotes equality.

We next compared SNP-level significance from the unadjusted test with that from covariate-adjusted regression using Groupwise variance estimates (Figure 5). For variants with MAF below 0.01, the distribution appeared more balanced around the identity line. In the remaining MAF strata, regions of higher variant density were predominantly above the identity line, indicating larger *−*log_10_(*p*) values and therefore smaller p-values after adjustment. A similar pattern was observed using HC1 variance estimates (Figure S1). This pattern is consistent with Wang et al.’s observation that the unadjusted sdMAF test can be conservative in samples comprising multiple populations,owing to the use of a population-pooled HWD estimate in variance estimation [1].

**Figure 5:**
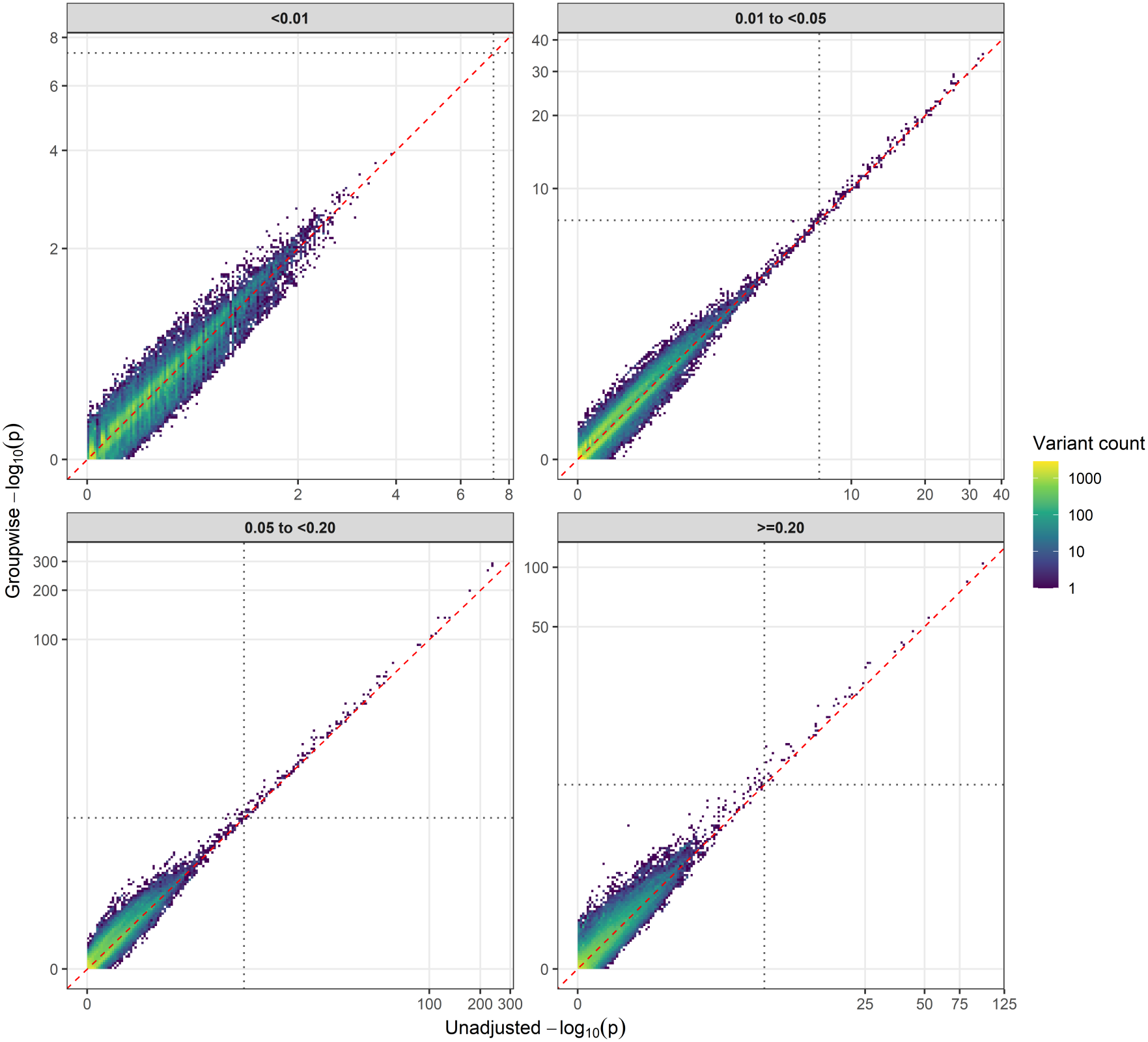
Comparison of association significance between unadjusted sdMAF and covariate-adjusted Groupwise tests. Two-dimensional binned density plots compare *−*log_10_(*p*) from the unadjusted test (horizontal axis) and adjusted regression with Groupwise variance estimates (vertical axis). Panels correspond to four MAF strata: *<* 0.01, 0.01-*<* 0.05, 0.05-*<* 0.20, and *≥*0.20. Both axes use a log1p transformation. Color indicates the number of variants per bin. The dashed red line denotes equality, and dotted lines indicate *p* = 5 *×* 10^*−*8^.

At the genome-wide significance threshold of *p <* 5 *×*10^*−*8^, 22 variants that were nonsignificant in the unadjusted analysis became significant with both HC1 and Groupwise variance estimates. Four additional variants gained significance with HC1 only, and one with Groupwise only, yielding 26 and 23 gained variants, respectively. Six variants significant in the unadjusted analysis were no longer significant with either adjusted variance estimator. Individual-variant estimates and significance results are provided in Table S1.

### 3.4 Sensitivity to variance estimation

The QQ plots showed that, in the upper tail, the unadjusted test generally produced smaller *−*log_10_(*p*) values than the three covariate-adjusted tests (Figure S2). This pattern is consistent with the previously reported conservativeness of unadjusted sdMAF testing in multi-population samples [1], although real-data QQ plots alone do not establish null calibration. To examine the contribution of variance estimation to differences among the adjusted tests, we compared their estimated variances directly (Figure 6).

**Figure 6:**
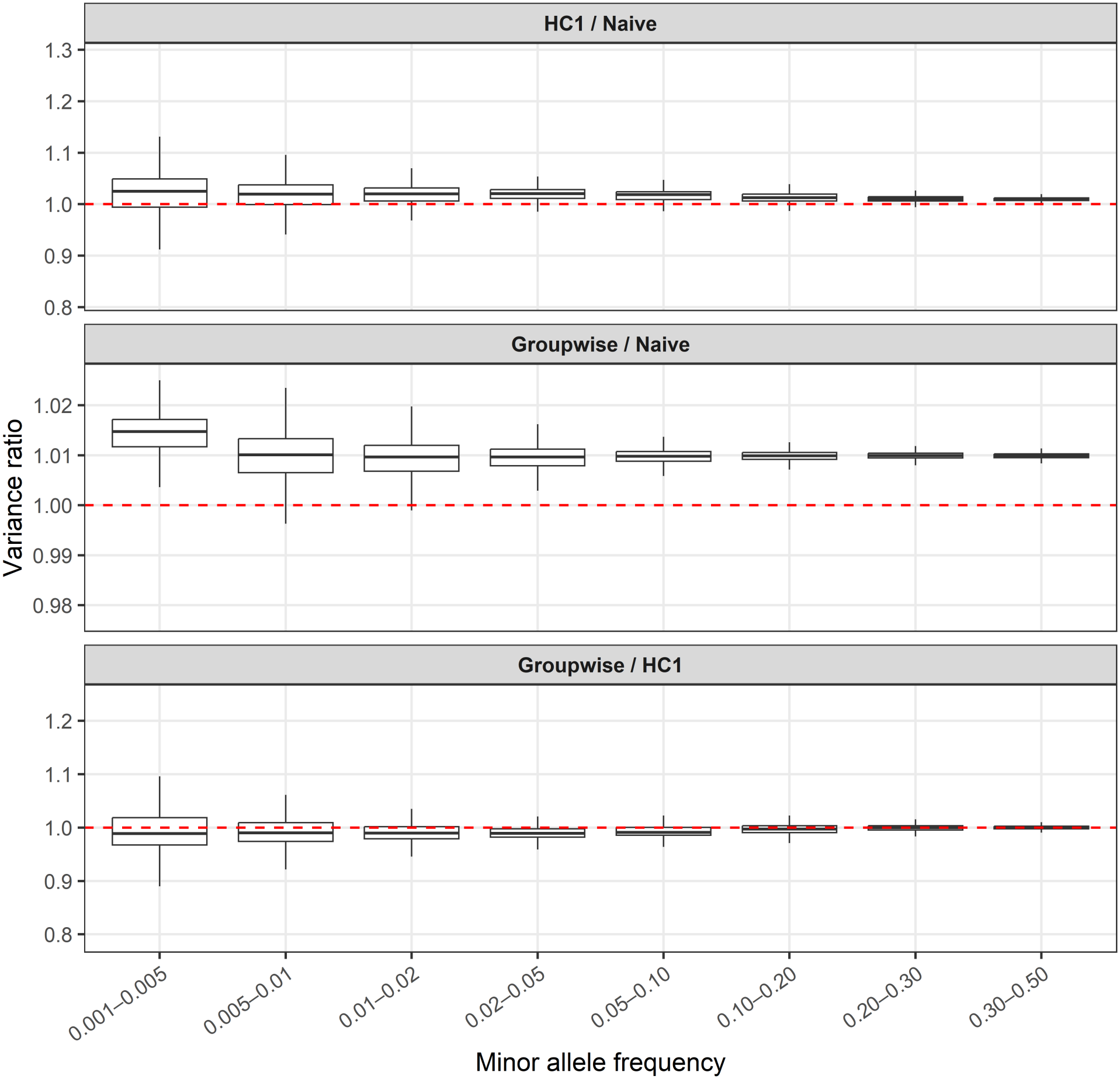
Comparison of variance estimates across MAF strata. Boxplots show HC1/Naive, Groupwise/Naive, and Groupwise/HC1 variance ratios from covariate-adjusted retrospective regression, stratified into nine MAF categories. The central line indicates the median, boxes represent the interquartile range, and whiskers extend to the most extreme observations within 1.5 interquartile ranges of the box. Outliers are omitted from visualization. The dashed red horizontal line denotes a variance ratio of one. All comparisons use the same allele-harmonized SNP set and identical adjusted point estimates.

Overall, HC1 and Groupwise produced larger variance estimates than the Naive estimator. The HC1/Naive and Groupwise/Naive ratios showed greater dispersion among variants with MAF below 0.01, whereas their distributions became more concentrated as MAF increased. In contrast, Groupwise/HC1 ratios were generally close to one. Their medians were slightly below one in the lower-MAF strata, indicating somewhat smaller Groupwise variance estimates relative to HC1, while agreement was closer in the higher-MAF strata.

The corresponding p-value comparisons showed a similar MAF-dependent pattern (Figure S3). For variants with MAF below 0.01, the binned densities were more dispersed around the identity line. As MAF increased, the densities became more closely concentrated along this line, indicating stronger concordance between HC1 and Groupwise inference. Together, these comparisons suggest broadly comparable performance of the two variance estimators in this dataset, with greater differences among lower-frequency variants.

## 4 Discussion

### 4.1 Main findings

We developed a regression-based framework for covariate-adjusted sdMAF inference in the X chromosome non-pseudoautosomal region (NPR). The framework provides adjusted subgroup estimates and their joint covariance, allowing inference on prespecified allele-frequency contrasts. In the scenarios examined, the Groupwise variance estimator performed similarly to HC1, whereas the Naive estimator was sensitive to sex sample imbalance. The unadjusted test frequently rejected the simulation null defined by the adjusted sex effect, illustrating the importance of distinguishing marginal and covariate-adjusted targets. The 1000 Genomes application further showed that adjustment can change SNP-level estimates and significance despite broad agreement between adjusted and unadjusted results.

### 4.2 Interpretation and relation to existing methods

Covariate-adjusted and marginal sdMAF address different questions. Marginal sdMAF describes the allele-frequency difference between the sampled female and male groups, including differences associated with their covariate distributions. The adjusted sex coefficient instead represents the contrast under the specified regression model. Neither target is inherently preferable in every setting; the choice depends on the scientific question and the role of the covariates.

Wang et al.’s omnibus test evaluates the joint null of no sdMAF in any population, whereas our primary application evaluates a single sex contrast adjusted for measured covariates. These hypotheses need not agree: population-specific sex differences may be present even when a specified overall contrast is zero. Their regression already accommodates sex- and population-specific variances. Our work makes the estimation and joint covariance of adjusted subgroup means explicit when nuisance coefficients are estimated from the full sample [5].

The distinction from the RA framework concerns both the inferential target and the treatment of nuisance covariates. Zhang and Sun’s autosomal method derives a score test for allelic association, whereas the subsequent X-chromosomal framework primarily addresses HWE and its relationship with sdMAF [4, 6]. Our analysis instead estimates adjusted subgroup allele-frequency contrasts and uses their covariance for Wald inference. The contribution is not the use of a Wald statistic itself, but the explicit covariance treatment and evaluation of residual-variance specifications for covariate-adjusted X-NPR sdMAF.

### 4.3 Variance estimation and its assumptions

Under male 0/2 coding, hemizygous male and diploid female genotypes have different variance-mean relationships. For example, at the same allele frequency under female HWE, male genotype variance is twice female genotype variance. A common residual variance can therefore be inappropriate.

The simple two-group comparison provides intuition for the Naive estimator’s sensitivity to sample imbalance. The pooled variance is weighted toward the larger group. When the smaller group has the larger variance, this can underestimate the variance of the group difference and inflate rejection rates. The reverse arrangement can produce conservative inference. Equal group sizes remove this discrepancy in the simple comparison, but do not generally guarantee validity after covariate adjustment, where the regression weights also matter. These considerations help explain the sex-ratio-dependent patterns observed in our simulations.

Groupwise estimation retains separate residual variance estimates for the prespecified groups. Its close agreement with HC1 in our analyses supports its use in the settings examined, but does not establish general equivalence. Groupwise assumes constant conditional residual variance within each group, whereas HC1 allows individual-level heteroskedasticity [9, 10]. Our simulations already permit within-sex variance variation through covariate-dependent allele frequencies, but do not exhaust the structures that could challenge Groupwise estimation.

### 4.4 Implications of the 1000 Genomes application

Adjustment for genetic principal components changed both sdMAF estimates and significance for individual variants. The tendency toward smaller adjusted p-values is consistent with Wang et al.’s observation that unadjusted sdMAF testing can be conservative in multi-population samples because population-pooled HWD estimates affect variance estimation [1]. However, our adjusted and unadjusted comparisons differ in both point estimates and estimated uncertainty, so the observed changes cannot be attributed solely to this mechanism.

The variance comparisons provided a more direct assessment of the adjusted estimators because Naive, HC1, and Groupwise use identical coefficient estimates. HC1 and Groupwise generally produced larger variances than Naive and agreed more closely with each other, particularly at higher MAF. Differences were greater among lower-frequency variants. Changes in genome-wide significance demonstrate the practical consequences of adjustment and variance selection, but do not establish which individual findings represent technical artifacts or biological differences.

### 4.5 Limitations

The adjusted contrast depends on the conditional mean model. HC1 can accommodate heteroskedasticity, but cannot recover the intended conditional contrast when relevant mean-model features are omitted. Groupwise estimation imposes additional variance assumptions whose adequacy should be assessed for the study population.

Both HC1 and Groupwise showed departures from nominal rejection rates in particular low-MAF scenarios, indicating finite-sample limitations. Our simulations used a nominal significance level of 0.05 with 10,000 replications per scenario. They therefore do not establish calibration at the much smaller thresholds used in genome-wide analyses. Further evaluation should examine extremetail behavior, a broader range of within-group variance structures, and sensitivity to mean-model misspecification. The derivations also assume independent observations; related samples require an appropriate covariance specification.

### 4.6 Extensions and conclusion

The joint covariance formulation allows adjusted subgroup estimates to be combined through prespecified linear contrasts. One application is the covariate-adjusted SPADE extension, in which sex-specific prevalence weights combine case and control estimates [13]. This extension is currently theoretical and requires empirical evaluation, including careful definition of the covariate reference distribution and consideration of uncertainty in prevalence estimates.

X-chromosomal association analysis requires careful treatment of genotype coding and dosage compensation [14]. For the allele-frequency estimand considered here, coding male genotypes as 0/2 places both sexes on the common scale E(*G*) = 2*p*, without imposing a dosage-compensation model for a disease phenotype. This rescaling does not equalize genotype variances. Groupwise or HC1 variance estimation addresses the remaining variance differences under its respective assumptions.

Allelic comparisons between sexes also have applications within disease cohorts. Mendes et al. used a case-only sdMAF analysis in an X-chromosome-wide study of ASD, illustrating the relevance of these comparisons beyond QC [15]. Extending covariate-adjusted inference to such applications requires specifying the target contrast, which should not be interpreted automatically as a logistic-scale gene-sex interaction.

The present empirical evidence concerns covariate-adjusted sdMAF in the X-NPR. Within this scope, the framework provides explicit estimation and inference, with Groupwise and HC1 yielding broadly comparable results across the settings examined.

## Data Availability

The human genetic data analyzed in this study are publicly available from the 1000 Genomes Project at https://ftp.1000genomes.ebi.ac.uk/vol1/ftp/data_collections/1000G_2504_high_coverage/working/20220422_3202_phased_SNV_INDEL_SV/. Genetic ancestry principal components are available in the PCs_1000G dataset in the PCAmatchR R package at https://cran.r-project.org/package=PCAmatchR. Simulation and analysis outputs are available from the corresponding author upon reasonable request.

https://cran.r-project.org/package=PCAmatchR

## Data availability

The high-coverage 1000 Genomes Project phased genotype data used in this study are publicly available from the 20220422 release at https://ftp.1000genomes.ebi.ac.uk/vol1/ftp/data_collections/1000G_2504_high_coverage/working/20220422_3202_phased_SNV_INDEL_SV/. Analyses were restricted to 2,504 individuals. The top 20 genetic ancestry principal components for these individuals were obtained from the PCs_1000G dataset in the PCAmatchR R package, available from CRAN at https://cran.r-project.org/package=PCAmatchR.

## Supplementary Materials

**Figure S1:**
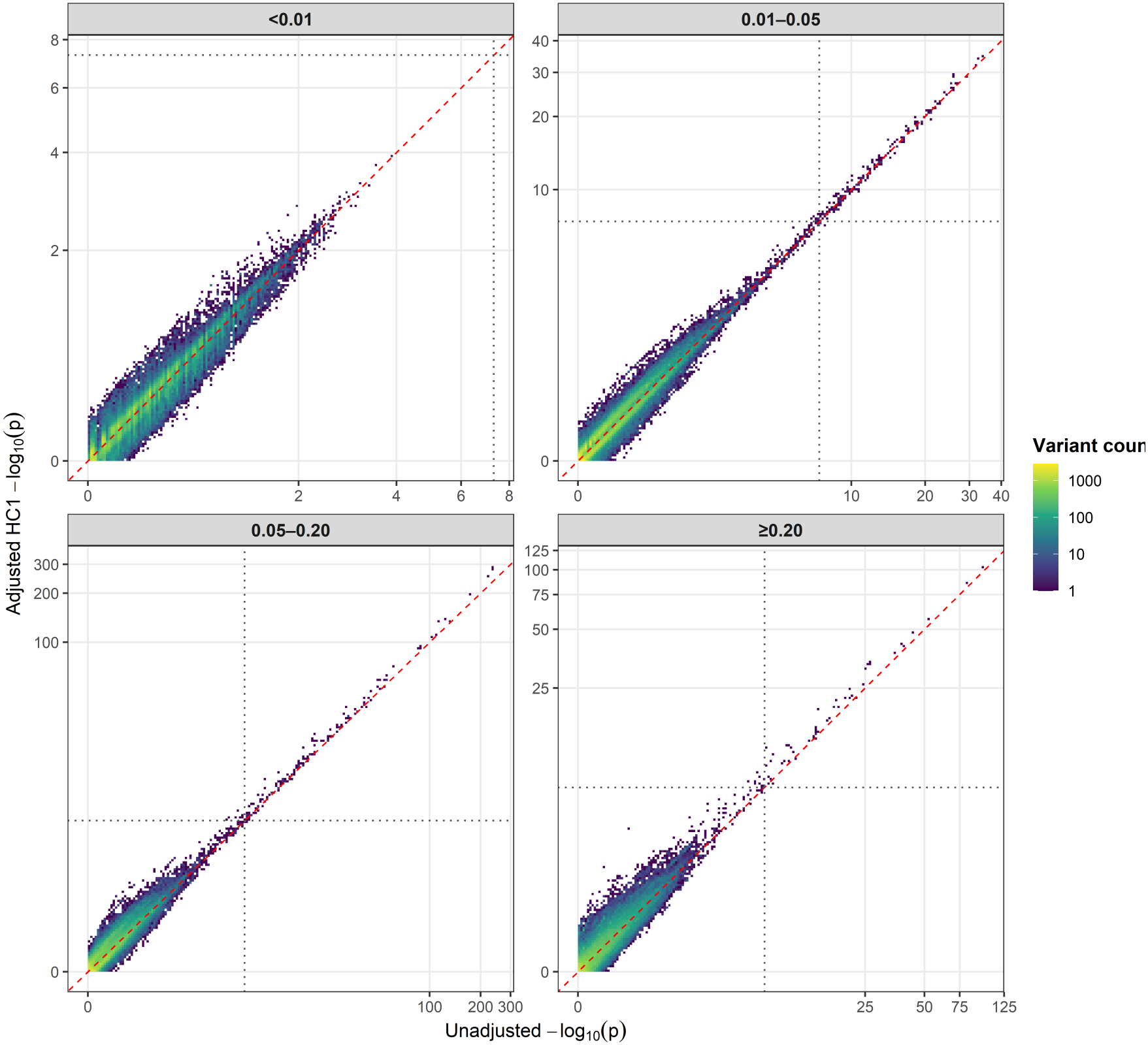
Comparison of association significance between unadjusted sdMAF and covariate-adjusted HC1 tests. Two-dimensional binned density plots compare *−*log_10_(*p*) from the unadjusted test (horizontal axis) and adjusted regression with HC1 variance estimates (vertical axis). Panels correspond to four MAF strata: *<* 0.01, 0.01-*<* 0.05, 0.05-*<* 0.20, and *≥*0.20. Both axes use a log1p transformation. Color indicates the number of variants per bin. The dashed red line denotes equality, and dotted lines indicate *p* = 5 *×* 10^*−*8^.

**Figure S2:**
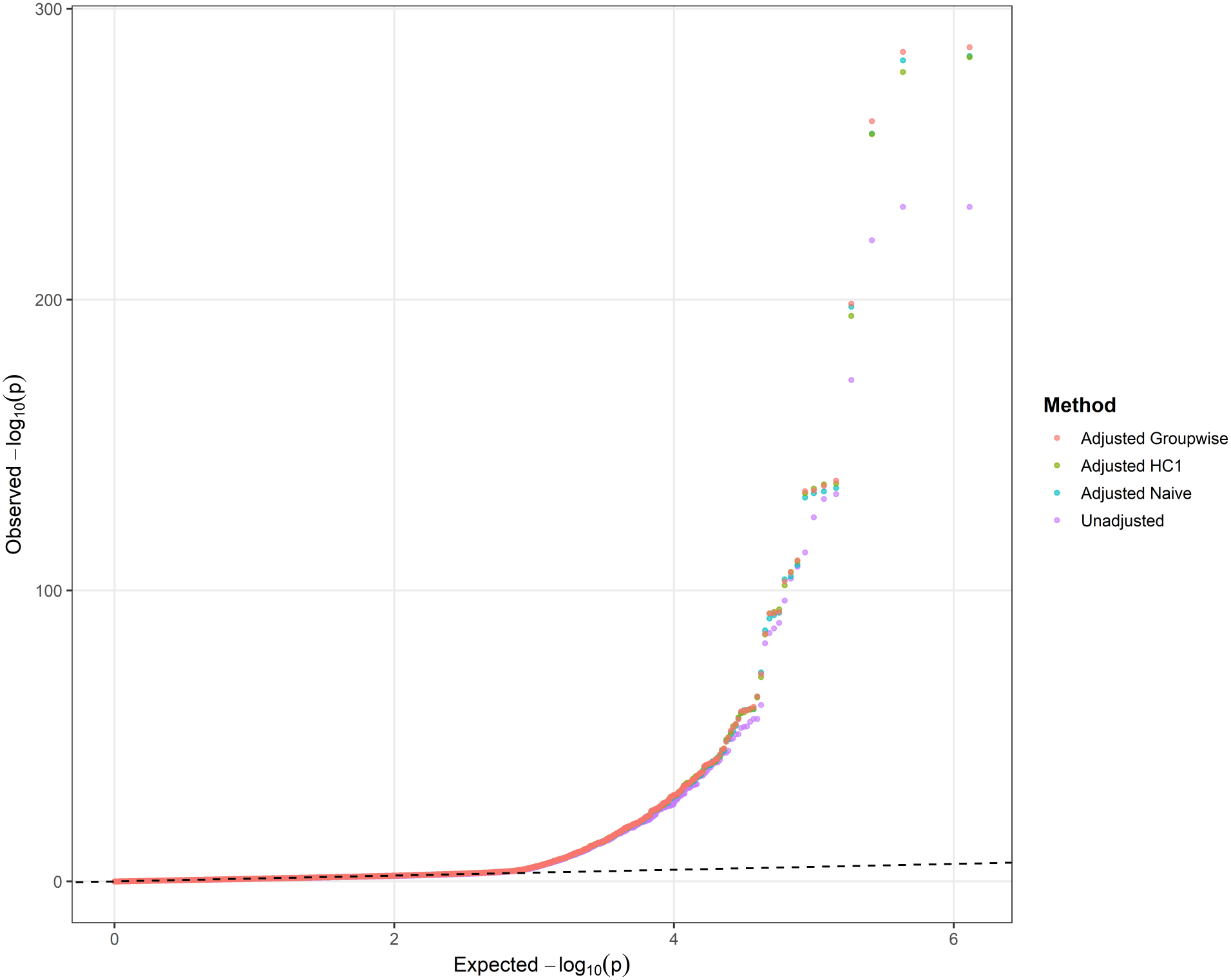
Quantile-quantile plots of unadjusted and covariate-adjusted sdMAF tests. Observed and expected *−* log_10_(*p*) values are compared for the unadjusted test and adjusted regression using Naive, HC1, and Groupwise variance estimators. Analyses use the same allele-harmonized SNP set. The dashed diagonal line represents the expectation under uniformly distributed null p-values. Departures may reflect genuine allele-frequency differences, residual confounding, or test miscalibration.

**Figure S3:**
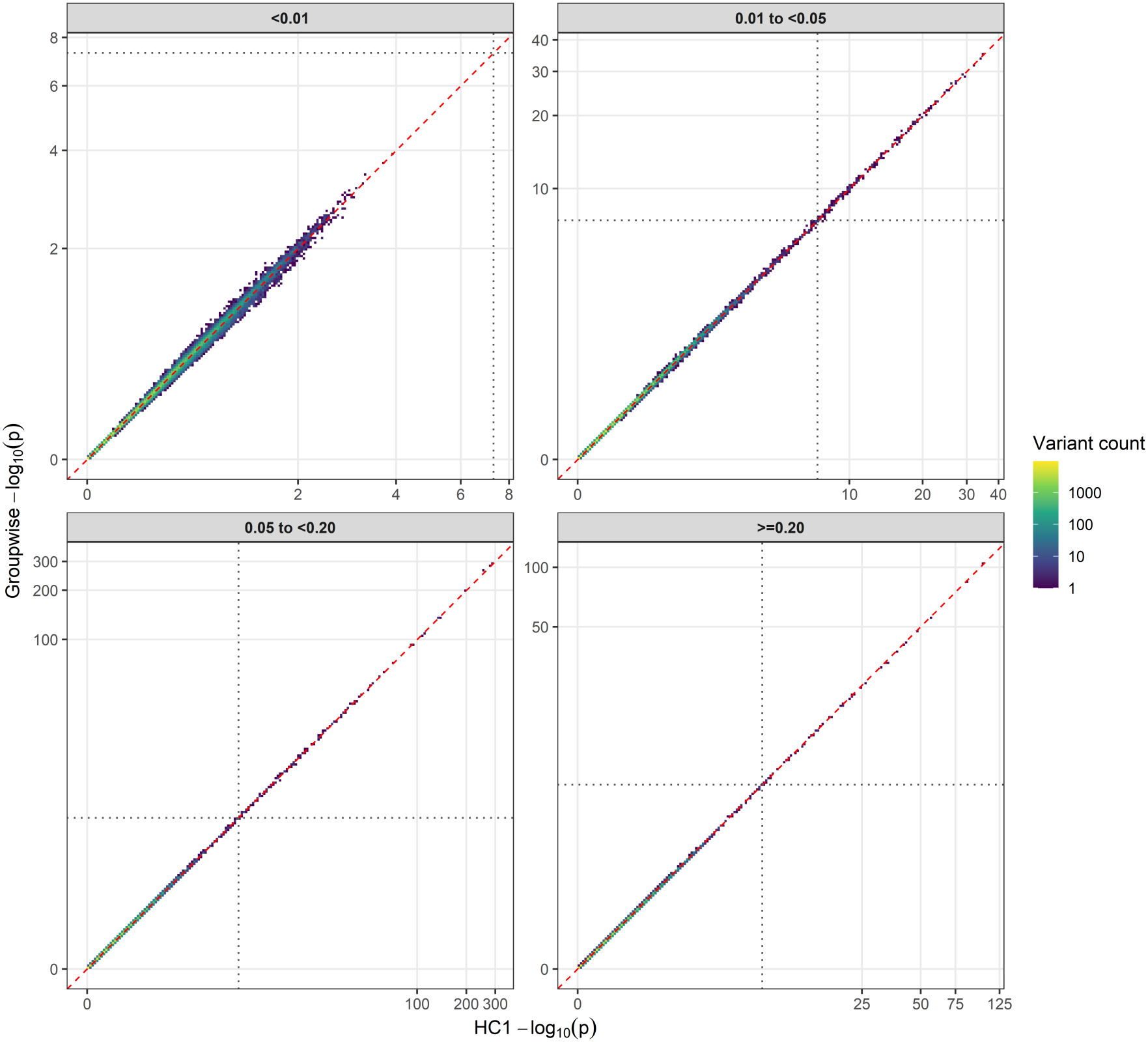
Comparison of association significance using HC1 and Groupwise variance estimates. Two-dimensional binned density plots compare *−* log_10_(*p*) from adjusted regression using HC1 (horizontal axis) and Groupwise (vertical axis). Panels correspond to four MAF strata: *<* 0.01, 0.01-*<* 0.05, 0.05-*<* 0.20, and *≥* 0.20. Both axes use a log1p transformation. Color indicates the number of variants per bin. The dashed red line denotes equality, and dotted lines indicate *p* = 5 *×*10^*−*8^. Both methods use identical adjusted point estimates; differences in significance arise from variance estimation.

**Table S1:** Variants gaining or losing genome-wide significance after covariate adjustment.

|  |  |  | sdMAF estimate |  | −log<sub>10</sub>(*p*) |  |  |
|---|---|---|---|---|---|---|---|
| Position on X | REF/ALT | MAF | Unadjusted | Adjusted | Unadjusted | HC1 | Groupwise |
| **A. Gained significance in at least one adjusted test** (*n* = 27) |  |  |  |  |  |  |  |
| 140,012,775 | G/T | 0.4214 | -0.0867 | -0.0807 | 6.596 | 8.839 | 8.854 |
| 120,083,314 | C/G | 0.1412 | 0.0559 | 0.0595 | 6.556 | 8.540 | 8.403 |
| 49,326,643 | G/A | 0.4533 | 0.0833 | 0.0897 | 6.857 | 8.329 | 8.375 |
| 9,410,806 | G/A | 0.2159 | 0.0725 | 0.0759 | 7.190 | 8.056 | 7.995 |
| 3,543,830 | A/C | 0.1664 | 0.0640 | 0.0669 | 7.154 | 8.046 | 7.910 |
| 102,463,352 | C/T | 0.2218 | 0.0655 | 0.0673 | 6.376 | 8.018 | 7.918 |
| 69,661,316 | A/G | 0.2582 | -0.0712 | -0.0760 | 5.892 | 7.955 | 7.896 |
| 6,534,588 | A/G | 0.0476 | 0.0343 | 0.0354 | 7.190 | 7.908 | 7.797 |
| 140,219,426 | T/G | 0.0763 | 0.0430 | 0.0416 | 6.872 | 7.820 | 7.842 |
| 153,316,077 | G/A | 0.1659 | 0.0588 | 0.0637 | 6.341 | 7.829 | 7.656 |
| 152,781,469 | C/A | 0.2050 | 0.0660 | 0.0697 | 6.819 | 7.785 | 7.685 |
| 149,544,091 | T/A | 0.0606 | 0.0391 | 0.0380 | 6.983 | 7.690 | 7.751 |
| 154,239,826 | A/C | 0.1021 | 0.0450 | 0.0429 | 5.623 | 7.593 | 7.698 |
| 72,162,064<sup>†</sup> | T/C | 0.0205 | 0.0207 | 0.0212 | 6.880 | 7.613 | 7.270 |
| 19,878,507 | A/T | 0.0412 | 0.0321 | 0.0311 | 7.187 | 7.505 | 7.584 |
| 144,107,143 | A/G | 0.4565 | -0.0858 | -0.0829 | 6.360 | 7.575 | 7.552 |
| 141,691,843 | G/A | 0.0221 | 0.0218 | 0.0225 | 7.040 | 7.529 | 7.483 |
| 32,807,150 | C/A | 0.0564 | 0.0380 | 0.0374 | 7.279 | 7.527 | 7.511 |
| 30,602,789 | G/A | 0.0194 | 0.0207 | 0.0203 | 7.297 | 7.500 | 7.396 |
| 136,441,150 | A/T | 0.3653 | -0.0848 | -0.0838 | 7.090 | 7.467 | 7.487 |
| 45,041,387 | A/C | 0.0189 | 0.0195 | 0.0202 | 6.735 | 7.481 | 7.392 |
| 125,152,249 | T/C | 0.0354 | -0.0294 | -0.0289 | 7.133 | 7.420 | 7.443 |
| 72,754,432<sup>†</sup> | G/C | 0.1949 | 0.0603 | 0.0644 | 5.526 | 7.400 | 7.286 |
| 120,067,941 | T/G | 0.2156 | 0.0717 | 0.0711 | 7.244 | 7.357 | 7.395 |
| 60,652,859<sup>†</sup> | G/C | 0.0191 | 0.0203 | 0.0199 | 7.108 | 7.393 | 7.270 |
| 69,910,386<sup>‡</sup> | T/A | 0.0484 | 0.0350 | 0.0344 | 7.253 | 7.218 | 7.316 |
| 7,593,103<sup>†</sup> | C/G | 0.0356 | 0.0298 | 0.0296 | 7.210 | 7.312 | 7.259 |
| **B. Lost significance in both adjusted tests** (*n* = 6) |  |  |  |  |  |  |  |
| 99,108,326 | C/A | 0.0173 | 0.0195 | 0.0187 | 7.705 | 7.085 | 7.107 |
| 116,557,764 | A/G | 0.0191 | -0.0331 | -0.0324 | 7.467 | 7.202 | 7.200 |
| 141,047,855 | T/G | 0.1239 | 0.0568 | 0.0538 | 7.448 | 7.063 | 7.101 |
| 112,023,246 | G/T | 0.0388 | 0.0309 | 0.0304 | 7.444 | 7.207 | 7.209 |
| 141,578,688 | A/G | 0.0739 | 0.0435 | 0.0427 | 7.364 | 7.116 | 7.127 |
| 119,475,584 | T/A | 0.0229 | 0.0230 | 0.0228 | 7.338 | 7.170 | 7.202 |

Genome-wide significance was defined as *p* < 5 × 10<sup>−8</sup>. Of 27 gained variants, 22 were significant with both estimators, four with HC1 only (†), and one with Groupwise only (‡), giving totals of 26 and 23, respectively. Six variants lost significance with both estimators. HC1 and Groupwise share adjusted estimates but differ in variance estimation. MAF, minor allele frequency; REF/ALT, reference/alternate alleles. Estimates are on the allele-frequency scale.

